# Diurnal changes and topographical distribution of ocular surface epithelial dendritic cells in humans, and repeatability of density and morphology assessment

**DOI:** 10.1101/2022.11.23.22282555

**Authors:** Zahra Tajbakhsh, Isabelle Jalbert, Fiona Stapleton, Nancy Briggs, Blanka Golebiowski

## Abstract

**Purpose:** Dendritic cells (DC) play a crucial role in ocular surface defence. DC can be visualised *in vivo* by confocal microscopy but have not yet been fully characterised in humans. This study investigated the diurnal variation, topographical distribution, and repeatability of DC density and morphology measurement.

**Methods:** *In vivo* confocal microscopy was conducted on 20 healthy participants (mean age 32.7±6.4 years, 50% F) at baseline and repeated after 30 minutes, 2, 6, and 24 hours. Images were captured at the corneal centre, inferior whorl, corneal periphery, limbus, and bulbar conjunctiva. DC density was counted manually, and morphology of DC was assessed for the largest cell body size, presence of dendrites, presence of long dendrites, and presence of thick dendrites. Mixed model analysis, non-parametric analyses, Bland & Altman plots, the Coefficient of Repeatability (CoR), and kappa were used.

**Results:** There were no significant changes in DC density (p≥0.74) or morphology (p>0.07) at any location over the 24-hour period. Highest DC density was observed at the corneal limbus followed by the peripheral cornea (p<0.001), with lowest density at the corneal centre, inferior whorl, and bulbar conjunctiva. Most DC at the corneal periphery, limbus, and bulbar conjunctiva had larger cell bodies compared to the corneal centre (p≤0.01), and presence of long dendrites was observed mostly at non-central locations. DC with thick dendrites were mostly observed at the limbus. Day-to-day CoR for DC density ranged from ±28.1 cells/mm^2^ at the corneal centre to ±56.4 cells/mm^2^ at the limbus. Day-to-day agreement of DC morphology determined by kappa ranged from 0.5 to 0.95 for cell body size, 0.60 to 0.95 for presence of dendrites, and 0.55 to 0.80 for presence of long dendrites, at various locations.

**Conclusions:** No diurnal changes are apparent in corneal or conjunctival DC. Substantial topographical differences exist in DC density and morphology. *In vivo* confocal microscopy provides good repeatability of DC density and acceptable agreement of DC morphology.

**Key points:**

- There was no diurnal variation in dendritic cell density, morphology, or topographical distribution at the ocular surface in healthy individuals.
- A gradient from high density of dendritic cells at the limbus to low density at the corneal centre was observed in *in vivo* confocal microscope images of healthy corneas. Density was lowest in the bulbar conjunctiva.
- The morphology of dendritic cells at the corneal periphery, limbus and bulbar conjunctiva suggests a greater antigen capture capacity compared to dendritic cells at the corneal centre.

## Introduction

Dendritic cells (DC) are a subtype of immune cells which play a pivotal role in the maintenance of ocular surface health and homeostasis and reside in the human cornea and conjunctiva.(1, 2) DC capture and process antigens and migrate to lymphoid organs through lymphatic vessels in the direction of cytokine gradients, where they present antigens to T cells for the initiation of an immune response.(3) DC have a specific morphology with a distinctive dendritiform shape.(4) Using *in vivo* confocal microscopy (IVCM), it is possible to examine DC in real time.

Observation of DC can be used to investigate changes occurring during ocular surface disease and to monitor the efficacy of treatment. Increases in DC density signal inflammation or upregulation of the immune response.(5) Changes in DC morphology can also be a marker of immune response activation, indicating the antigen capture and migratory capacity of these cells as summarised and reviewed.(2, 6-19) Ocular surface DC present in a steady state in ocular tissues that are in contact with the external environment and, in the event of an injury or insult, they undergo significant alteration to activate the immune response.(2) Upon activation, DC undergo morphological changes induced by the expression of surface markers such as major histocompatibility complex.(3) Various changes in DC morphology including increased cell size, cell body size, number, length, thickness, field area and cell area of dendrites, have been reported during corneal inflammation.(6, 10, 18, 20) These changes can reflect the antigen capture and migratory capacity of DC.(15)

Evaluation of alterations in DC density and morphology in the context of ocular surface disease or treatment, requires the use of repeated observations. Characterisation of variations in DC density and morphology over the course of a day, and day-to-day repeatability of measurements is thus essential to enable normal variations to be differentiated from changes that can be attributed to disease processes. This knowledge of DC variations would also help facilitate the effective design for future studies, including sample size determination, in the studies assessing ocular surface DC.

Diurnal changes in ocular surface DC density or morphology have not yet fully been investigated. The circadian rhythm is a key component of immune cell function. Daily oscillations in lymphocyte trafficking and migration, including circulating blood DC, are an indication that they are under circadian control.(21) Higher circulating leukocyte counts were found during the day in mice and at night in humans based on their rest-activity times.(22-24) Daily fluctuations in number of lymphocytes were observed in blood, lymph, and lymph nodes in an animal model.(25, 26) A recent study reported no significant changes in corneal DC density during the sleep/ wake cycle.(27) However, the density of DC with long dendrites was higher at the corneal centre upon awakening (04:30 and 06:30) compared to the midday visit (between 09:00 to 12:00).(27)

Day-to-day repeatability of corneal DC density or morphology has not previously been reported. A strong correlation between two central corneal DC density measurements taken 3 weeks apart using IVCM has been reported in normal patients.(28) Other corneal locations have not yet been investigated. Day-to-day repeatability of bulbar conjunctival DC density has been reported to be ±5.3 cells/mm^2^ in a PhD thesis.(29)

DC have been observed in many corneal and conjunctival locations, but to date, their topographical distribution (density or morphology) has not been thoroughly examined in a single IVCM study. A high limbal DC density and a low central corneal density have been consistently reported, as well as a low DC density at the bulbar conjunctiva.(30-33) DC with longer dendrites and a larger dendritic field area have been observed at the corneal periphery compare to the centre in humans and animals.(18, 31, 34, 35)

This study aimed to evaluate diurnal changes in corneal and conjunctival epithelial DC density and morphology over a 24-hour period and to determine day-to-day repeatability of those measurements. A secondary aim was to explore topographical differences in DC density and morphology at various ocular surface locations.

## Methods

A prospective, observational, investigator-masked study was conducted. DC density and morphology were measured at 5 timepoints over the course of 24 hours and at 5 ocular surface locations. The study was approved by the Human Research Ethics Committee of UNSW Sydney. The study followed the Declaration of Helsinki, and informed consent was obtained from all participants prior to the start of the study.

### Participants

Twenty participants from UNSW Sydney were recruited from the September 2019 to January 2020. Participants were aged 18 years or older and had healthy ocular structures. Exclusion criteria included active ocular surface disease and intraocular inflammation, systemic or ocular disease likely to affect the ocular surface, systemic or ocular allergy, ocular surgery or injury (reported by the participants or observed on ocular examination), current use of topical medications including ocular lubricants, antihistamines, systemic anti-inflammatory medications, pregnancy or breastfeeding, regular soft contact lens wear, or any history of rigid contact lens wear. Sample size calculation (GPower 3.1) showed that 20 participants were sufficient to detect a DC density change of 10 cells/mm^2^ between repeats with 80% power and a confidence level of 95% (assuming a standard deviation of 15 cells/mm^2^, correlation between repeated measures of r=0.60, and an adjustment of 15% for non-parametric data).(36)

### Clinical procedures

IVCM was conducted at baseline (3 hours after waking and before 11am) and after 30 minutes, 2, 6, and 24 hours. A Rostock Corneal Module attached to an HRT III IVCM (Heidelberg Engineering GmbH, Heidelberg, Germany) was used to visualise DC. At each timepoint, IVCM was conducted on the right eye of all participants at the following 5 topographical locations, in the following order: corneal anatomical centre, corneal inferior whorl, far peripheral cornea (temporal, 1mm inside limbus), limbal cornea (temporal), and bulbar conjunctiva (temporal, 2-3 mm from the limbus) (Figure 1). Unpreserved topical anaesthetic (oxybuprocaine hydrochloride 0.4%, Minims, Bausch & Lomb UK) was instilled in the right eye and GenTeal gel (hypromellose 0.3%, carbomer 980 0.2%, Ciba Vision Ophthalmics, Australia) was used as a coupling medium between the TomoCap, a sterile and transparent cap, and the objective lens. The focus was changed manually to image the level of the corneal sub-basal nerve plexus and conjunctival epithelium, and a set of images (400 × 400 µm) were captured using section mode.

**Figure 1:**
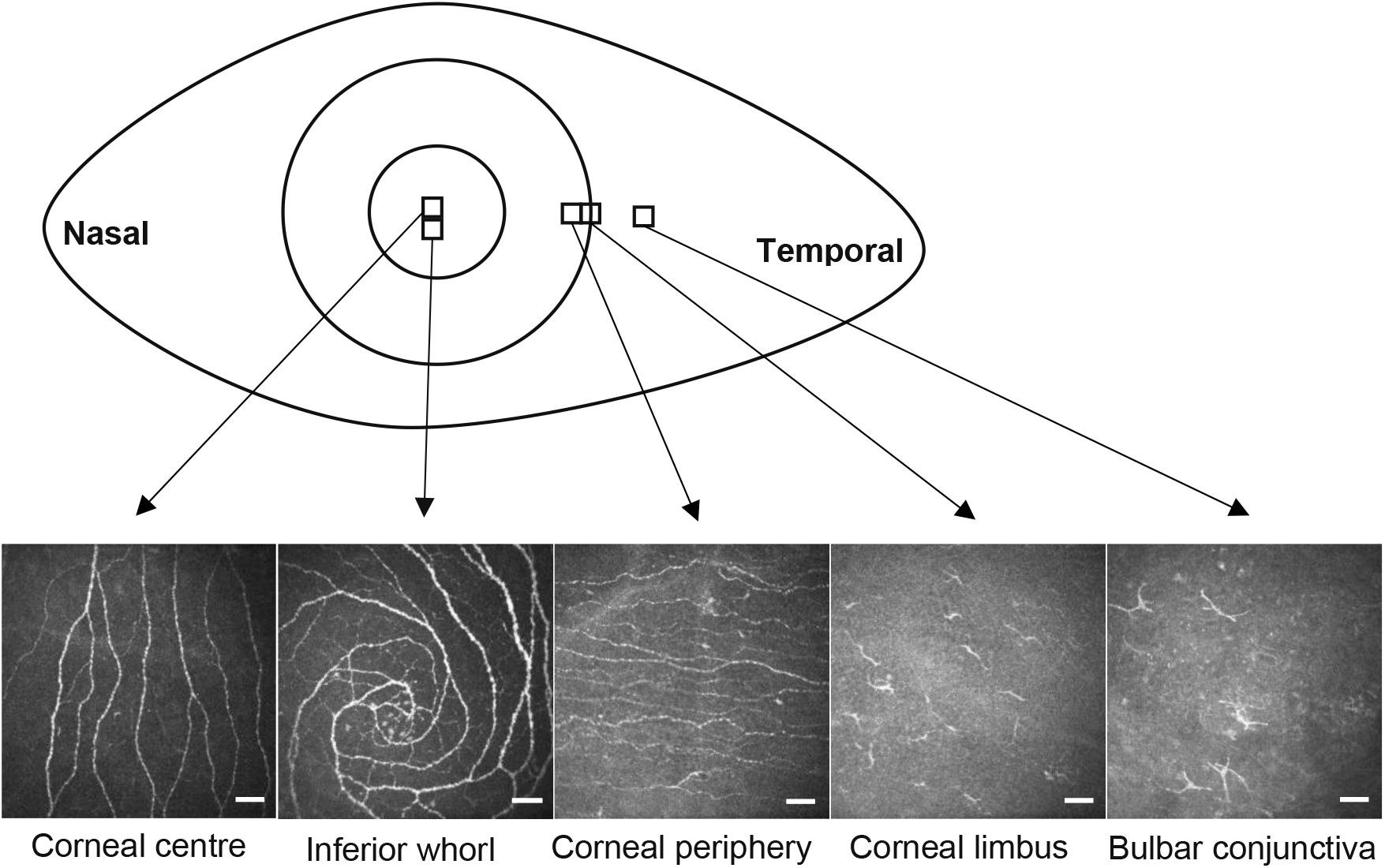
Schematic of the five topographical locations examined by *in vivo* confocal microscopy. Representative images (400 × 400 µm) are provided from each ocular surface location. Bars indicate 50 µm.

### Image analysis

Approximately 200 digital images were captured from all ocular surface locations for each participant. For each of the topographical locations, 5 images were selected for analysis, based on those which were best focused and overlapping by less than 20%. Only one single best focused image was selected from the inferior whorl due to its small area. Bright/ hyperreflective cells at least 10 µm in size, with a linear/ curvilinear cell body, with or without dendrites (both short and long), located at the subbasal corneal epithelium and distributed among nerve fibres that do or do not cross them or conjunctival epithelium were considered as DC.(15) To minimise bias, DC density and morphology were assessed by an experienced investigator who was masked to the participants’ identity and measurement timepoint.

#### DC density

The number of any cell based on the above definition for DC in each image was counted manually. The mean number of DC in the 5 images is reported for corneal centre, periphery and limbus, and conjunctiva. The number of DC in a single image is reported for the inferior whorl.

#### DC morphology

DC body size and presence of dendrites at all locations were assessed using a grading system developed by our group.(15) Cell body size refers to the cell soma and does not include the dendrites and was graded as small (10-25 µm), medium (26-40 µm), or large (>40 µm), based on the largest cell body size observed in any of the 5 images (or in the single inferior whorl image). The measurement was based on an estimation of cell body size by using a pictorial reference as described previously by our group without further analysis by software to make it easy to use and accessible.(15) The presence of any dendrites, the presence of long dendrites and the presence of thick dendrites in any of the 5 images (or in the single inferior whorl image) were recorded. Images devoid of DC were not included in the DC morphology analysis.

### Statistical analysis

Data analysis was performed using SAS (SAS 9.4 (2012); SAS Institute Inc., Cary, NC, USA)) and SPSS software (version 26; SPSS Inc.). Differences in DC density between timepoints and between locations were examined using a linear mixed model. Density data were log-transformed after adding 0.5 to density values of zero before entering into the mixed model. Differences in DC cell body size between timepoints and between corneal locations were examined using a generalized linear mixed model with multinomial distribution. For both models, fixed effects of timepoint, location and their interaction were included, and a random effect for individual was included to account for the repeated measures within person. Pairwise differences between locations at each timepoint were obtained within each model, and p-values adjusted using Holm’s stepdown Bonferroni method. Due to a small number of DC observed in conjunctival images, differences in conjunctival cell body size between timepoints were assessed using the Friedman test and comparisons with corneal locations were assessed using the Wilcoxon Signed-Rank test and p-values adjusted using Holm’s stepdown Bonferroni method. The number of data points was also small for presence of dendrites at corneal and conjunctival locations, hence differences in presence of dendrite appearance between timepoints and between locations were assessed using the Cochran’s Q test.

Repeatability of day-to-day measurements (baseline and 24h) of DC density was assessed using Bland and Altman plots and the coefficient of repeatability (CoR) was calculated. For DC morphology, Cohen’s weighted kappa was used to assess agreement between day-to-day measurements (baseline and 24h) for cell body size, and kappa was used for presence of dendrites.(37) In cases where high agreement was observed, but kappa / weighted kappa values were low (referred to as the “kappa paradox”), Gwet’s AC1 was used instead.(38) Associations between DC density at various locations were assessed at baseline using Pearson correlation. P-values less than 0.05 were considered statistically significant.

## Results

Twenty participants (mean age 32.7±6.4 years, 50% female) with healthy ocular surface, and no report of ocular surface disease as per inclusion criteria completed the study.

### DC Density

DC were observed in all participants at all corneal locations, other than at the inferior whorl, where no DC were evident in 5 to 7 participants at various timepoints. Conjunctival DC were observed for 13 to 15 participants at various timepoints. The linear mixed model analysis showed no significant effect of time, over 24 hours, on DC density (F=0.01, p=0.99) and no significant interaction between timepoint and location (F=0.11, p=1.0) showing that location did not impact this finding. A significant effect of location on DC density was evident (F=137.09, p<0.001). Simple main effects to examine the DC changes over the 24-hour period within each location were also assessed and are presented in Table 1 and Figure 2.

**Table 1:** Median (interquartile range) dendritic cell density (cells/mm^2^) at the ocular surface over a 24-hour period using *in vivo* confocal microscopy in 20 participants with healthy eyes.

| Timepoint<br>Location | DC density cells/mm<sup>2</sup> |  |  |  |  | p-value |
| --- | --- | --- | --- | --- | --- | --- |
|  | Baseline | 0.5 Hours | 2 Hours | 6 Hours | 24 Hours |  |
| Corneal centre | 11.9 (4.1-52.8) | 11.9 (2.8-57.2) | 11.2 (2.5-68.1) | 10.6 (2.8-52.8) | 11.9 (4.1-55.9) | 0.98 |
| Inferior whorl | 9.4 (0-46.9) | 12.5 (0-37.5) | 12.5 (1.6-34.4) | 12.5 (0-31.2) | 15.6 (0-31.2) | 0.99 |
| Corneal periphery | 35.0 (18.7-56.2) | 35.6 (19.1-44.4) | 33.7 (19.4-49.7) | 28.1 (14.1-57.5) | 39.4 (13.7-53.4) | 0.74 |
| Corneal limbus | 80.6 (51.6-113.7) | 85.0 (50.0-102.5) | 87.5 (48.7-107.5) | 82.5 (50.9-127.8) | 92.5 (56.6-101.9) | 0.99 |
| Bulbar conjunctiva | 3.7 (0-21.6) | 3.1 (0.3-20.3) | 3.7 (0-17.2) | 4.4 (0.3-20.3) | 6.9 (0.3-17.5) | 0.99 |
| p-value | < 0.001 | < 0.001 | < 0.001 | < 0.001 | < 0.001 |  |

**Figure 2:**
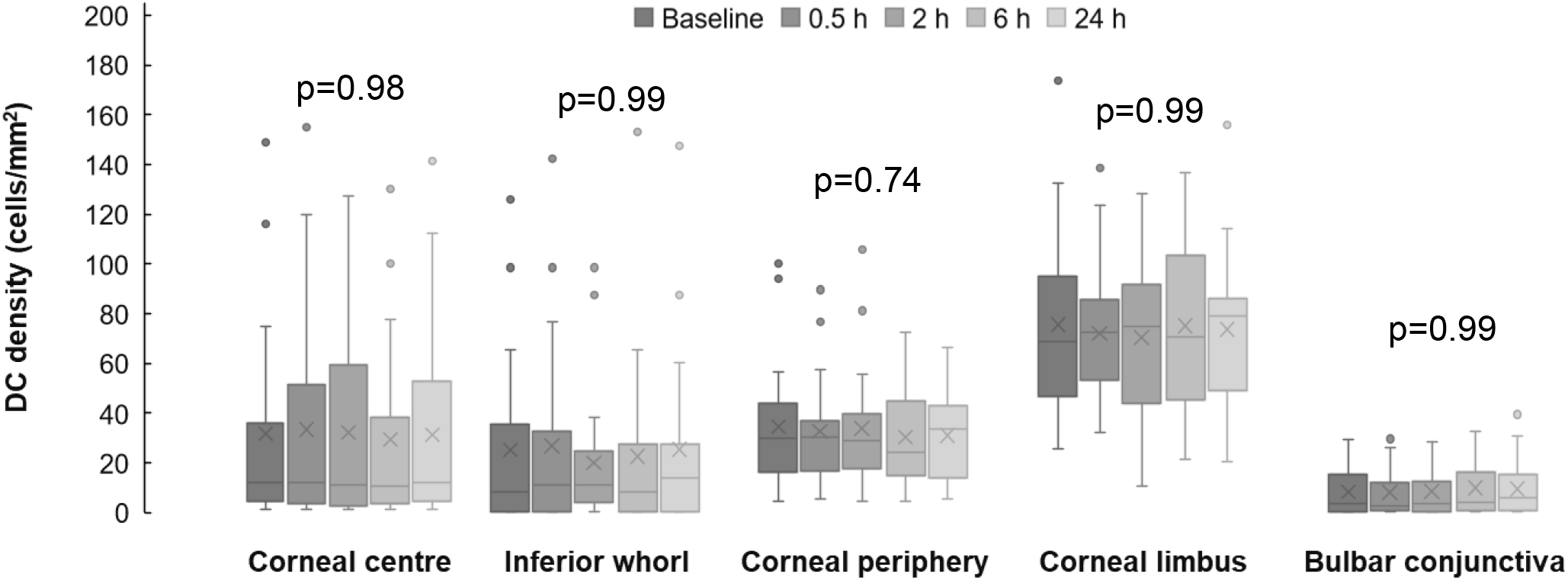
Diurnal variation of dendritic cell density at the ocular surface, observed over a 24-hour period using *in vivo* confocal microscopy, in 20 participants with healthy eyes. Plots represent median (horizontal grey line), mean (cross), interquartile range (box), lower and upper extremes (whiskers) and outliers lying above Q3+1.5*interquartile range and below Q1-1.5*interquartile range (circles).

At all timepoints, the highest DC density was observed at the corneal limbus (p<0.001) followed by the corneal periphery (p≤0.05). The lowest corneal DC density was at the centre and inferior whorl (p≤0.05) which were not significantly different (p=1.0). Differences between DC density at the corneal periphery and all other locations were significant (p≤0.03), other than the difference between the corneal periphery and centre at the baseline timepoint only (p=0.05). The lowest DC density was observed at the bulbar conjunctiva (p<0.001), which was not significantly different to the corneal centre or inferior whorl (p≥0.10).

Associations between DC density at various locations were examined at baseline. Higher DC density at the corneal centre was associated with higher DC density at the inferior whorl (r=0.67, p=0.001) and corneal periphery (r=0.66, p=0.001) but not with density at the corneal limbus. Higher DC density at the corneal limbus was associated with higher density at the corneal periphery only (r=0.57, p=0.01). DC density at the bulbar conjunctiva was not significantly associated with density in any of the corneal locations (r ≥ -0.16, p ≥ 0.51) (Supplementary Table 1 and Figure 1).

### Day-to-day repeatability of DC density

DC density measurements conducted at the same time of day on consecutive days (baseline and 24h) were not significantly different at any location (p≥0.21, Figure 3). Figure 3 shows the differences in DC density between baseline and 24h measurements on the y-axis and the mean of the two measurements on the x-axis, for each location. At the corneal centre, the CoR of ±28.1 cells/mm^2^ and the limits of agreement indicate that 95% of the differences between the two measurements can be expected to lie between +28.4 and -27.8 cells/mm^2^. The CoR was similar for the inferior whorl, values were relatively higher at the corneal periphery and limbus, and the smallest CoR was calculated for the conjunctiva (Figure 3).

**Figure 3:**
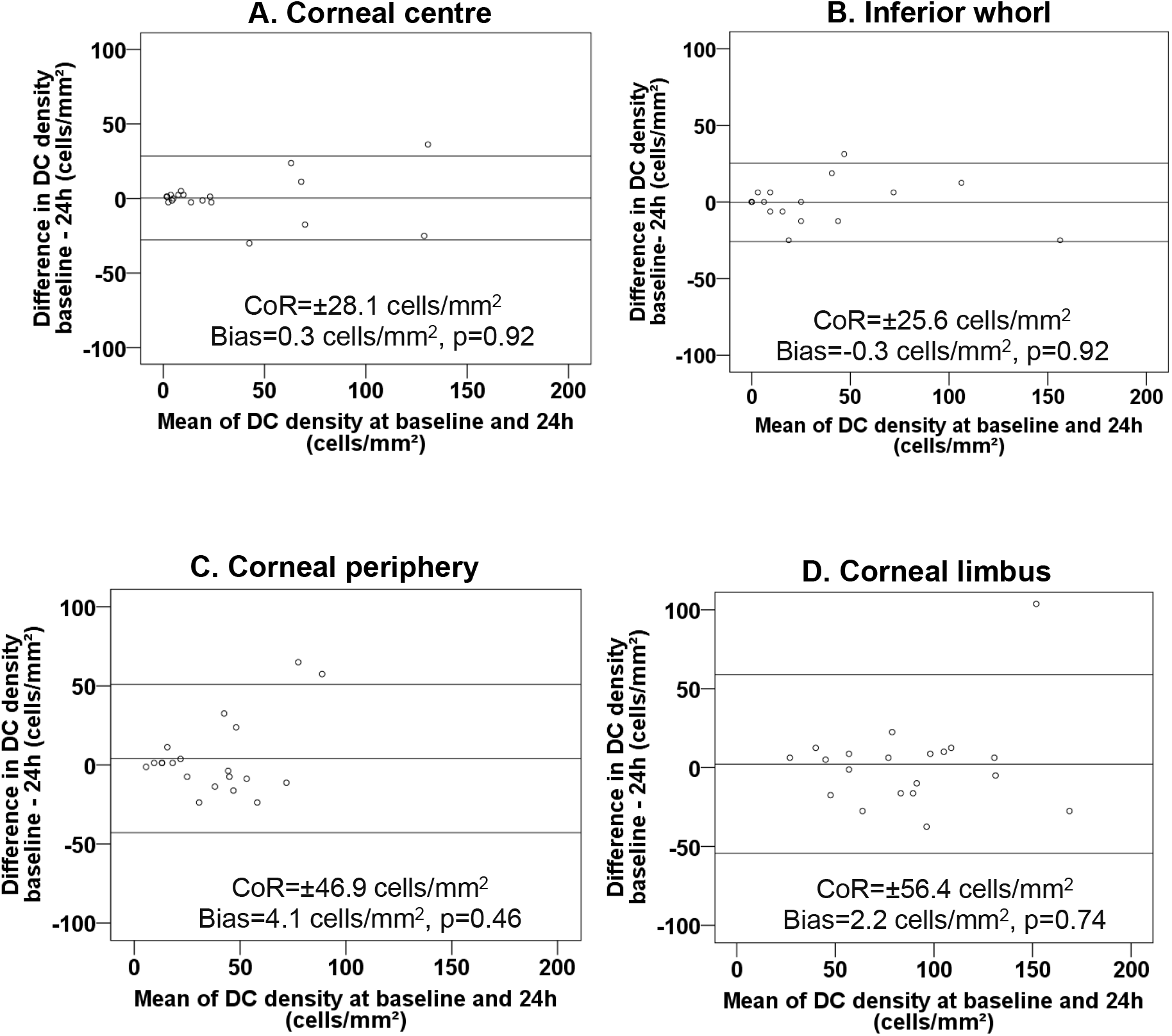
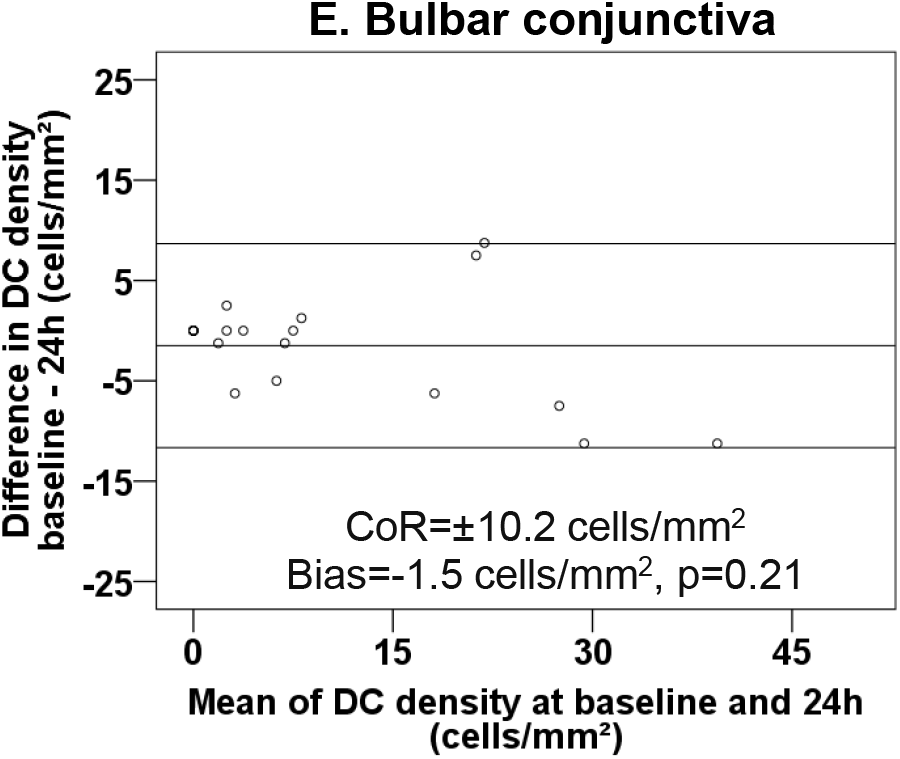
Ocular surface dendritic cell density measurements at 5 topographical locations in 20 participants with healthy eyes. The difference in density measurements obtained at two timepoints 24 hours apart is plotted against their mean at each corneal (A to D) and conjunctival (E) location. Horizontal lines indicate the bias (central line) and the limits of agreement (upper and lower lines).

### DC Morphology

DC morphology did not change significantly over the 24-hour period at any of the locations (p≥0.07). DC morphology data for the baseline timepoint only is presented in Figure 4; data for all other timepoints are provided in supplementary data (Supplementary Figure 2 A-E and Table 2 F-H).

**Figure 4:**
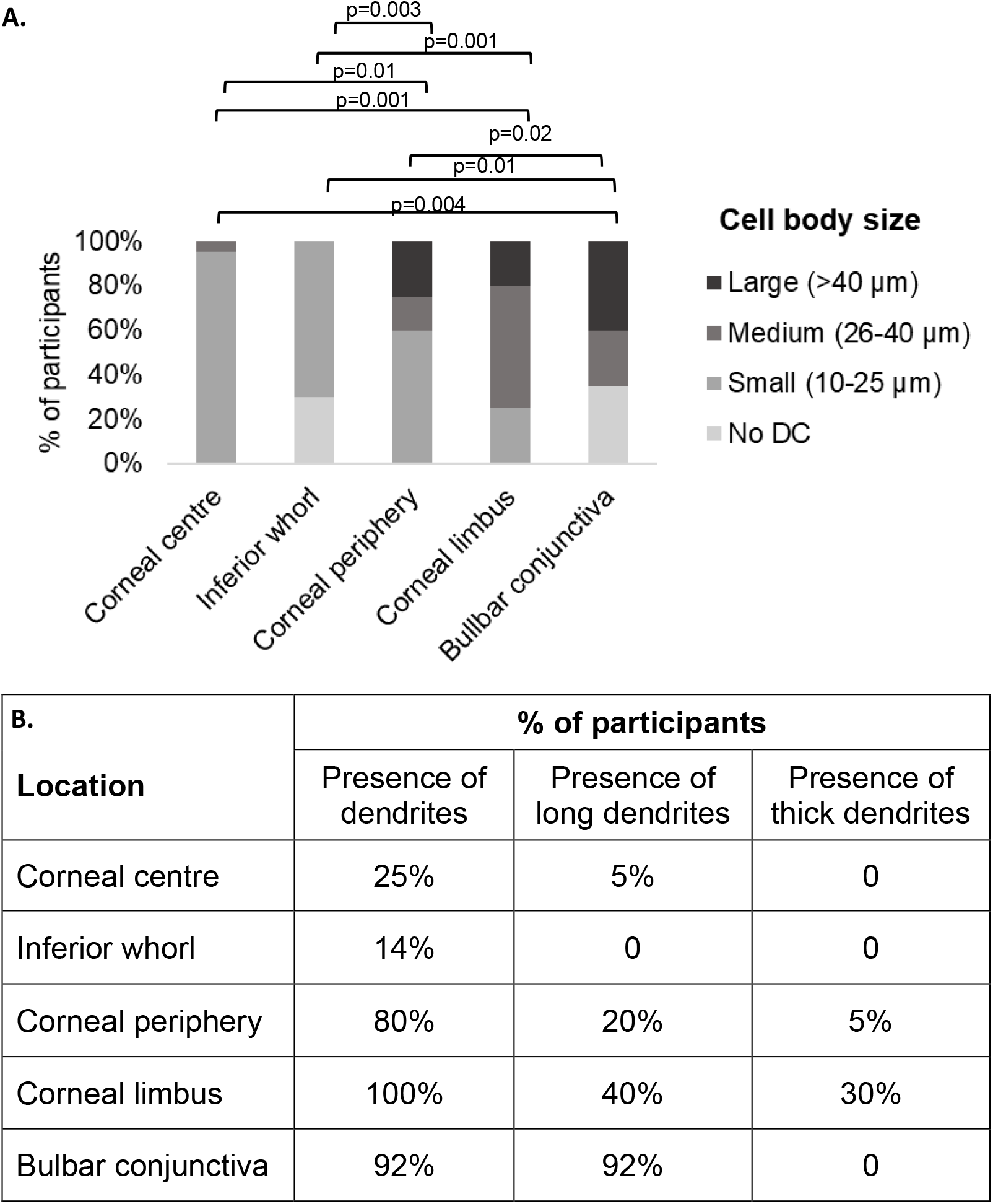
Ocular surface dendritic cell morphology at baseline: A. cell body size, B. dendrite appearance (presence of dendrites, presence of long dendrites and presence of thick dendrites) in 20 participants with healthy eyes. Images devoid of DC are not included in the table and analyses and thus data are presented for 14 participants only for the inferior whorl and 13 participants for the bulbar conjunctiva. No DC segment in the cell body size figure is for the reference only and was not used in the analyses.

**Table 2:** Agreement for ocular surface dendritic cell morphology measurements conducted on two consecutive days (baseline and 24 hours) in 20 participants with healthy eyes. NA: agreement was not assessed due to the low frequency of occurrence of long dendrites at the corneal centre, and of thick dendrites at any location.

|  | Degree of agreement |  |  |
| --- | --- | --- | --- |
|  | Cell body size<br>(Cohen's weighted<br>kappa) | Presence of<br>dendrites<br>(kappa) | Presence of long<br>dendrites<br>(kappa) |
| Corneal centre | 0.95* | 0.90* | NA |
| Corneal periphery | 0.50* | 0.60* | 0.55 |
| Corneal limbus | 0.70* | 0.95* | 0.80 |
| Bulbar conjunctiva | 0.50 | 0.85 | 0.60 |

\*Gwet's AC1 used in place of Cohen's weighted kappa / kappa.

At corneal locations, no significant effect of time on cell body size was evident (F=0.56, p=0.70). There was no interaction between timepoint and location (F=0.37, p=0.97), and a significant effect of location on cell body size was evident (F=72.88, p<0.001). A larger cell body size was observed at the corneal periphery and corneal limbus compared to the corneal centre and inferior whorl at all timepoints (p≤0.01). Cell body size was not significantly different between the corneal centre and inferior whorl (p=1.0), nor between the corneal periphery and limbus at any timepoint (p≥0.11). Almost all (94%) DC at the corneal centre and inferior whorl had a cell body size of less than 25µm.

Conjunctival DC body size also did not change significantly between timepoints (p=0.67). The morphology of DC observed at the bulbar conjunctiva was distinct from that of corneal DC (Figure 5). DC bodies at the conjunctiva were larger in size than those at the corneal centre or inferior whorl at all timepoints (p≤0.01), and larger than those at the corneal periphery at baseline and 24h (p≤0.03). The differences in cell body size between conjunctival and limbal DC were not significant at any timepoint (p≥0.10).

**Figure 5:**
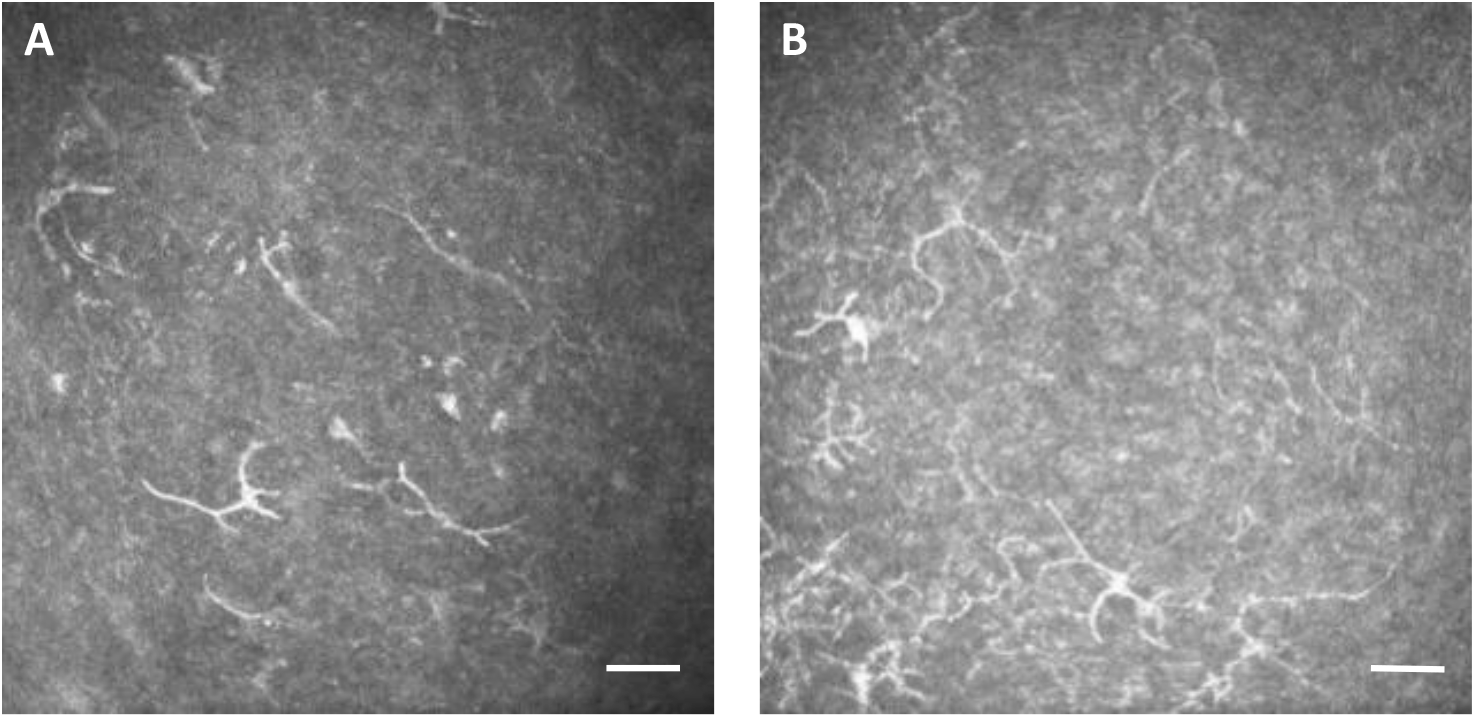
A and B: Representative *in vivo* confocal microscopy images of temporal bulbar conjunctival epithelial dendritic cells. In some cases, conjunctival DC were observed forming an X- and Y-shaped pattern (A), and sometimes forming a sheet or layer (B). Image size is 400 × 400 µm and scale bar indicates 50 µm.

There were no significant differences between timepoints in the proportion of DC with dendrites (p≥0.40), nor in the proportion of DC with long dendrites (p≥0.07), at any location (Supplementary Table 2 F and G). The proportion of DC with thick dendrites also did not appear to differ between timepoints but statistical analysis could not be conducted due to the low frequency of occurrence of thick dendrites, A greater proportion of DC with dendrites were observed at the corneal periphery, limbus and bulbar conjunctiva compared to the inferior whorl at all timepoints (p≤0.01) and more were observed at the limbus compared to the corneal centre at baseline, 2h and 24h (p≤0.04) (Supplementary Table 2 F-H). Almost all DC at the limbus were observed to have dendrites, as did most DC at the corneal periphery and at the conjunctiva (Figure 4B, Supplementary Table 2 F). In contrast, approximately one third of DC at the corneal centre had dendrites, and very few at the inferior whorl. The majority of conjunctival DC presented with long dendrites, and one third to half of DC at the corneal limbus and periphery. Very few DC at corneal centre had long dendrites, and none at the inferior whorl. Thick dendrites were observed only at the corneal limbus (approximately a quarter of DC) and very few at the corneal periphery.

### Day-to-day agreement of DC morphology

Agreement for DC cell body size between two timepoints 24 hours apart was moderate at the corneal periphery, limbus, and conjunctiva, with almost perfect agreement at the corneal centre (Table 2). Agreement was almost perfect for the presence of dendrites at the corneal centre, limbus, and conjunctiva and moderate at the corneal periphery. Agreement for the presence of long dendrites was moderate to perfect at corneal periphery, limbus, and bulbar conjunctiva. Due to the low frequency of occurrence of long dendrites at the corneal centre, and thick dendrites at any location, agreement was not assessed. Also, agreement was not assessed at the inferior whorl due to no variability of cell body size, low frequency of occurrence of dendrites and no occurrence of long or thick dendrites.

## Discussion

In this study for the first time, diurnal variation and repeatability of DC density and morphology measurements at various ocular surface locations were assessed and shown to be stable and repeatable. Topographical differences in DC density and morphology between multiple corneal and conjunctival locations are shown for the first time, with the highest density (likely reflecting highest antigen capture capacity) of DC found at the corneal limbus and the least density (and least antigen capture capacity) of DC found at the corneal centre. Defining of diurnal variation and repeatability is important to develop an understanding of the natural history of ocular surface conditions; this information is also essential in the monitoring of disease progression and the effectiveness of treatment.

No diurnal variation in DC density or morphology was observed at any ocular surface location in this study. Although there are no previous reports in the literature of diurnal variation of DC, the current results are aligned with those of the control arm of an 8 hour intervention study, which found no changes in central corneal DC density in 10 healthy participants.(39) There are no such observations for other topographical locations nor for DC morphology. In a mouse model, leukocyte trafficking to the corneal limbal region showed a circadian rhythm with a peak in neutrophil and γδ T-cell levels in the dark phase of the experiment.(40) Mice models in areas other than the eye have shown more lymphocytes and DC at night in the lymph node (22), and a reduction in the number of B cells, CD4^+^ and CD8^+^T cells in blood during the dark phase.(25, 26) Overall DC density at the corneal centre and inferior whorl region has shown to be constant during the sleep/wake cycle, with an increase in the number of DC with long dendrites at the corneal centre only after awakening.(27) Of note, assessment was limited in this study to a time frame ranging from three hours after awaking to a maximum of 5 PM and thus it is possible that DC density could vary during sleep and at night. Nevertheless, any changes in DC occurring during sleep are not likely to impact clinical assessments which are typically conducted during the daytime.

Day-to-day repeatability of DC density was shown to be moderate for the corneal centre (CoR = ±28.1), inferior whorl (CoR = ±25.6), bulbar conjunctiva (CoR = ±10.2), and relatively lower for the corneal periphery (CoR = ±46.9) and limbus (CoR = ±56.4). The larger CoR for DC density measurements at the corneal periphery and limbus is proportionate to the higher DC density observed at these two locations but may also reflect difficulties with returning to the same limbal and peripheral corneal location during IVCM image capture.(41) A size effect is evident in the Bland & Altman plots for corneal centre, periphery, and conjunctiva, indicating smaller limits of agreement at lower DC density values. The calculated CoRs may thus underestimate repeatability of DC density for values below approximately 40 cells/mm^2^ at the corneal centre, 30 cells/mm^2^ at the corneal periphery and 15 cells/mm^2^ at the conjunctiva. At the same time, the calculated CoRs may overestimate repeatability for DC density above these values, which might be of relevance for studies of highly inflamed eyes where higher DC density measurements might be expected.

Only two studies have reported on repeat measurements of corneal or conjunctival DC density. Central corneal DC density measurements at two visits, three weeks apart were shown to be highly correlated, with no significant difference in means between measurements; however, repeatability was not calculated.(28) The CoR for conjunctival DC density measurement is larger herein compared to that reported earlier in a PhD thesis (CoR: ±5.3 cells/mm^2^).(29) Methodological differences may account for this discrepancy, as well as the location of sampling (temporal versus nasal bulbar conjunctiva). The repeatability of DC density at other corneal locations including the corneal periphery and limbus has not been reported previously. The higher CoR values at these locations are not surprising as corneal curvature at the far peripheral cornea and limbus makes image capture more difficult, and thus it is more challenging to return to the exact same location in these locations for repeat measurements. This limiting factor in determining repeatability is due to the small field of view of IVCM and image sampling.(33, 42)

Repeatability of DC morphology has not previously been explored. The good day-to-day agreement of cell body size and presence of dendrites at the corneal centre and limbus obtained herein suggests that in healthy eyes DC morphology is stable and its measurement sufficiently repeatable to apply in clinical trials of disease progression and/or treatment monitoring. The low frequency of occurrence of thick dendrites and of long dendrites at some locations did not allow day-to-day agreement of dendrite appearance to be accurately calculated and this requires confirmation in a larger sample.

This study constitutes the first full topographical representation of all corneal and conjunctival locations measured concurrently *in vivo* in the human eye. In normal eyes without ocular surface disease, the lowest DC density in the cornea is observed at the centre, with highest density at the corneal limbus. The observed distribution agrees with previous work investigating specific locations. DC density in the normal human cornea has previously been reported to be highest at the limbus (range 33.5 to 288 cells/mm^2^), then at the periphery (range 37.9 to 108.8 cells/mm^2^) and lowest at the corneal centre (range 5.1 to 53 cells/mm^2^).(30, 31, 33, 43-45) A relatively lower DC density has been observed at the conjunctiva (range 7.0 to 42.1 cells/mm^2^).(32, 33, 46)

In this study DC density at the corneal centre was associated with density at the inferior whorl and corneal periphery, but not with the corneal limbus. This agrees with the strong association in DC density between the central and mid-peripheral cornea previously reported in a healthy group including contact lens and non-contact lens wearers.(47) The lack of association between DC density at the corneal centre and limbus could be due to the steep difference in baseline densities between these two locations, which might reflect independent recruitment of cells at each location. Corneal and conjunctival DC density were not associated with each other in this study, and this warrants further evaluation, for example to elucidate whether these corneal and conjunctival DC are the same DC subtypes. Heterogenous phenotypes of DC have been observed in the naïve conjunctiva of mice, including Langerhans cells, and myeloid DC.(48) Conventional DC are reported as the main resident DC in the cornea.(49)

Few studies have previously assessed the distribution of DC morphology at the ocular surface, and cell body size of DC in human corneas.(19) In line with the current findings, longer dendrites have previously been reported at the corneal periphery compared to the centre in healthy human corneas assessed using IVCM.(35) Similarly, in a study of participants with mild dry eye, there were more DC with long dendrites at the corneal periphery compared to the corneal centre where there were none.(50) Another IVCM study in normal human cornea described dendritiform cells with dendrites mainly at the periphery, and cells lacking dendrites at both central and peripheral cornea which is similar to that reported in animal studies.(2, 31, 34) In a study of healthy mice, a larger DC field area (area outlined by the dendrite process tips) was measured at the corneal periphery compared to the centre.(18)

In the current study, DC bodies were larger at conjunctiva than corneal locations and larger at corneal limbus and periphery than corneal centre and inferior whorl. Almost all DC at the corneal periphery had dendrites and a third had long dendrites. Most of the limbal DC had dendrites; a third had long dendrites and almost all conjunctival DC had long dendrites. DC with larger cell bodies and DC with long dendrites can be a marker of effective antigen capture capacity of DC.(7, 8, 10, 12, 14, 15, 31, 51-53) The findings of this study suggest that antigen capture capacity of DC in the corneal periphery and limbus is superior to that at the centre, and that of conjunctival DC is superior to corneal cells. The morphology of conjunctival DC observed in the current study is consistent with previous reports using IVCM.(54-56)

The presence of thick dendrites is an indicator of migratory capacity of DC.(9, 15, 57) DC with migratory potential were evident mostly at the limbus. This may be due to their close proximity to limbal vessels which facilitate DC migration.(58) This is the first study to describe the morphology of DC at the limbus.

DC observed at the corneal centre and inferior whorl had small cell bodies and few had dendrites which were short but not thick. This agrees with two earlier IVCM studies of healthy corneas, which observed small DC with short or no dendrites to be predominant in the corneal centre.(43, 44) These morphological features suggest these cells have low antigen capture and low migratory capacity, indicating that these are the least mature DC. Notably, it is possible that these cells are another type of immune cell, as DC these cells cannot easily be identified using IVCM due to the intrinsic limitation of IVCM by providing 2D images, and immunohistochemistry methods by co-staining with DC markers may reveal this.

In the current study, we used IVCM to assess human DC density and morphology at the corneal and conjunctival epithelium. The main intrinsic limitation of IVCM is that it cannot confirm the cell phenotype and provide the information about the cell surface markers to confirm the cells observed are in fact DC. Cross-validation of IVCM findings with immunohistochemistry methods which is possible in the conjunctiva, not human corneal tissue would be of benefit.

In conclusion, diurnal variation of DC density and morphology was not observed at the ocular surface. The good day-to-day repeatability of DC density measurements and good agreement of DC morphology grading provides researchers with a clear indication that any DC changes observed using IVCM can be used to monitor progression of ocular surface inflammation and to monitor treatment effectiveness. Differing DC density and morphology patterns observed at various corneal and conjunctival locations may indicate differences in immune function. The corneal periphery, limbus and bulbar conjunctiva appear to have DC with the most antigen capture capacity, whereas DC at the limbus have the most migratory capacity.

## Data Availability

All data produced in the present study are available upon reasonable request to the authors.

## Acknowledgments

This study was supported by the University of New South Wales (UNSW) and the Faculty of Science Research Infrastructure Scheme. The first author received a fee remission scholarship from UNSW Sydney and Australian Government Research Training Program Scholarship.

## Conflict of interest

No conflicting relationship exists for any author.

---

**Supplementary Figure 2 (A-E) and Table 2 (F-H):**
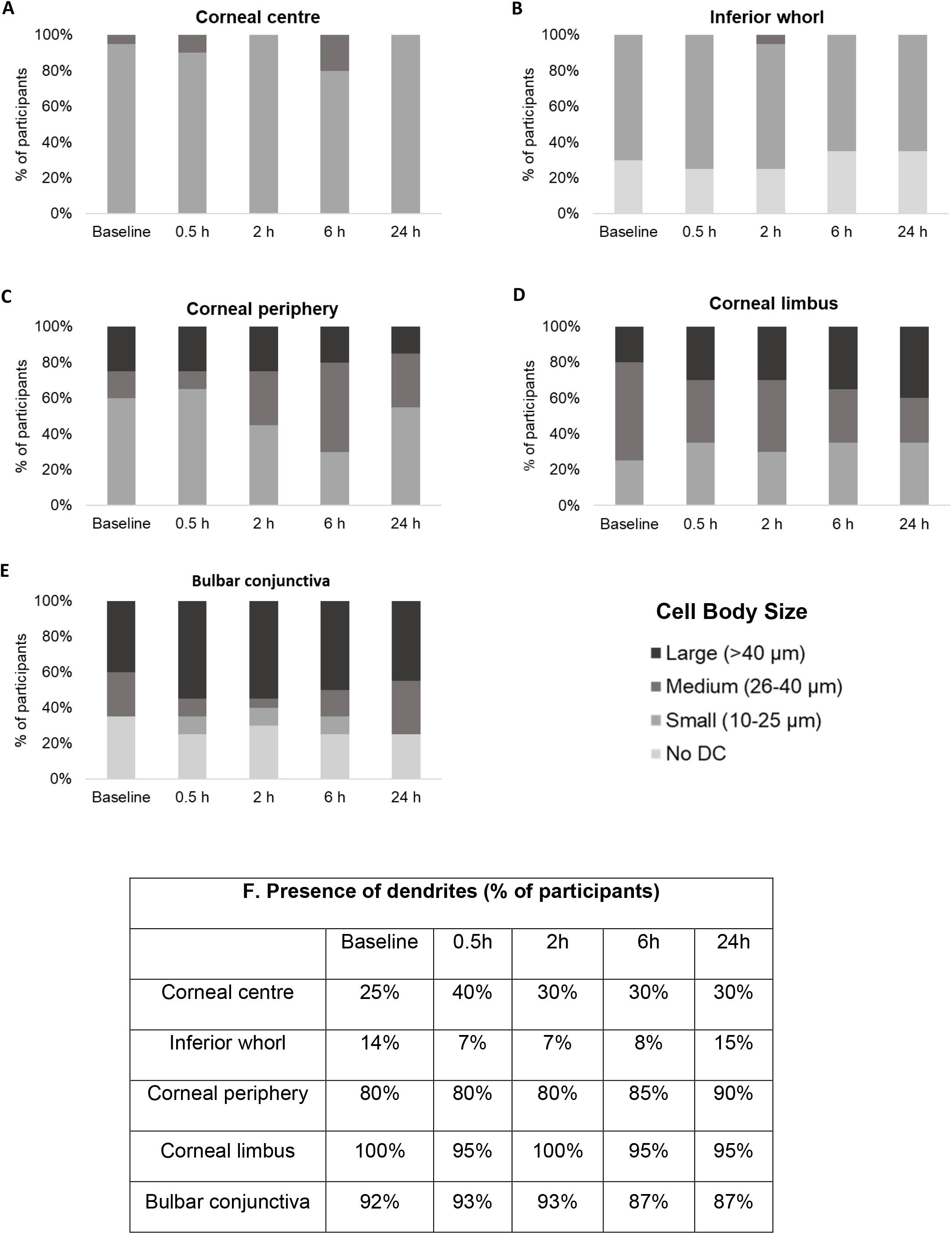
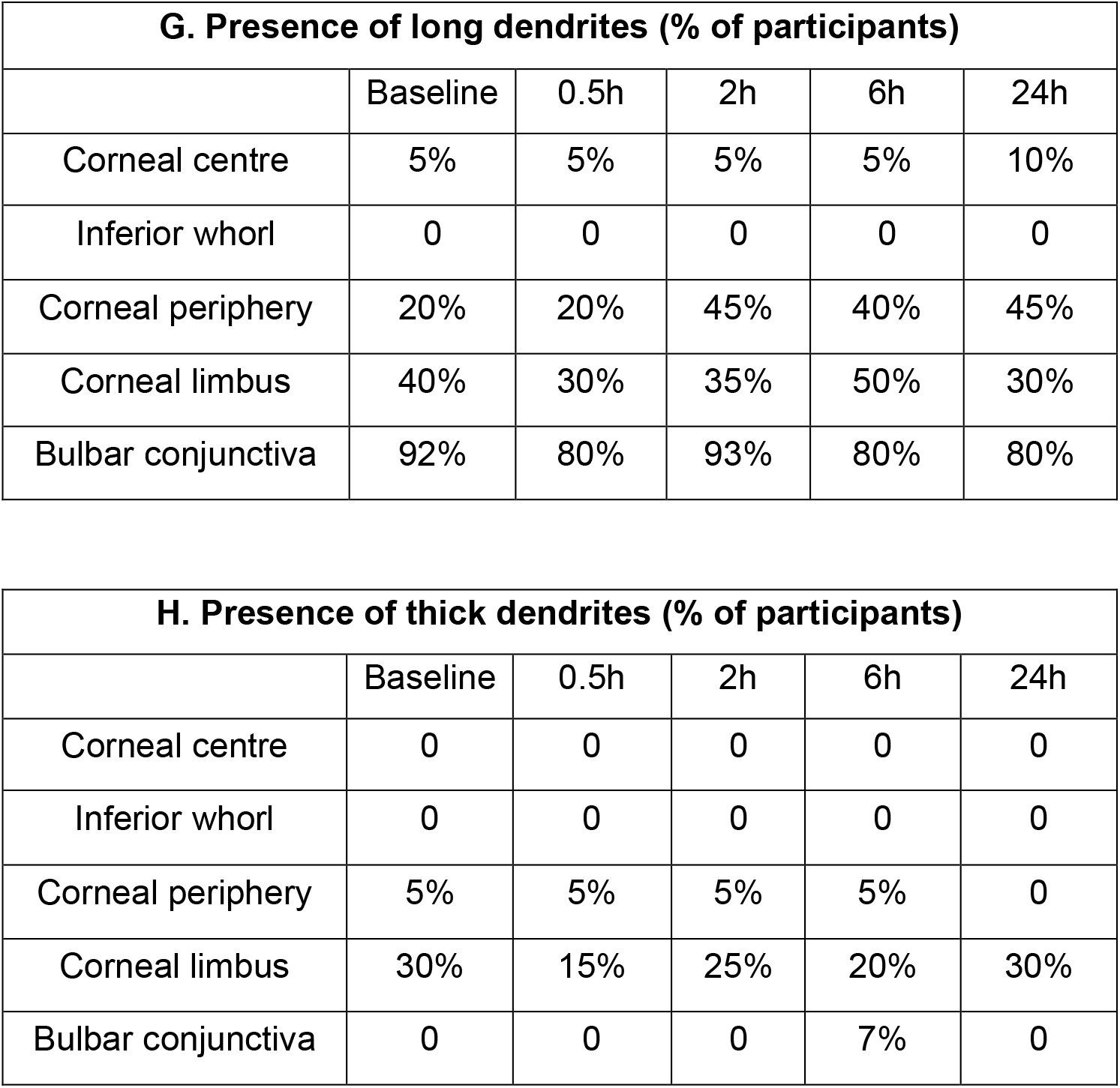
Diurnal variation of dendritic cell morphology over a 24-hour period at the ocular surface in 20 participants with healthy eyes. Four morphological parameters are presented: cell body size (bar charts A-E), presence of dendrites (Table F), presence of long dendrites (Table G) and presence of thick dendrites (Table H). Images devoid of DC are not included in the table and analyses and thus data are presented for only 13 to 15 participants for the inferior whorl and bulbar conjunctiva at different timepoints. No DC segment in the cell body size figure is for the reference only and was not used in the analyses.

**Supplementary Table 1 and Figure 1 (A-C):** Associations between dendritic cell density at ocular surface locations at baseline. Values are Pearson correlation, P-value. Significant correlations are in bold/italics. A graphical representation of the association between corneal centre and inferior whorl (Figure A), corneal periphery (Figure B), and between corneal periphery and corneal limbus (Figure C) is shown.

| Topographical Location | Inferior whorl | Corneal periphery | Corneal limbus | Bulbar conjunctiva |
| --- | --- | --- | --- | --- |
| Corneal centre | **0.67, 0.001** | **0.66, 0.001** | 0.27, 0.25 | 0.10, 0.65 |
| Inferior whorl | -- | **0.66, 0.001** | 0.37, 0.10 | -0.02, 0.94 |
| Corneal periphery | -- | -- | **0.57, 0.01** | -0.16, 0.51 |
| Corneal limbus | -- | -- | -- | -0.15, 0.53 |

## Notes

### Competing Interest Statement

The authors have declared no competing interest.

### Funding Statement

This study did not receive any funding.

### Author Declarations

The study was approved by the Human Research Ethics Committee of University of New South Wales Sydney.

